# Early Population-Level Impact of *Helicobacter pylori* Eradication on Gastric Cancer Deaths in Japan: A Counterfactual Analysis of Short-Term Divergence

**DOI:** 10.64898/2026.02.24.26346975

**Authors:** Akiko Kowada

**Author notes:** Correspondence to: Akiko Kowada.

## Abstract

**Background:** *Helicobacter pylori* infection accounts for 98% of gastric cancer (GC) cases in Japan. Since 2013, the nationwide expansion of *H. pylori* eradication therapy to chronic gastritis patients has created a unique opportunity to evaluate its population-level impact on GC primary prevention. However, short-term reductions in GC deaths are difficult to interpret given the long natural history of gastric carcinogenesis. This study aimed to assess the early impact of population-level eradication on GC deaths.

**Methods:** We applied a two-layer analytic framework consisting of a counterfactual analysis comparing observed GC deaths during 2013–2021 with expected GC deaths had eradication uptake remained at pre-2013 levels. This was combined with a structured, time-dependent, multilayer state-transition model to estimate GC deaths prevented by eradication using GC incidence integrated with age-dependent *H. pylori* prevalence.

**Results:** Observed GC deaths declined from 48,632 in 2013 to 41,624 in 2021, whereas counterfactual GC deaths declined more modestly, from 49,794 to 45,654. The divergence between observed and counterfactual GC deaths widened steadily from 1,162 in 2013 to 4,030 in 2021. Model-based estimates indicated that eradication prevented 1,427 GC deaths during 2013–2021, with annual GC deaths prevented increasing from 17 in 2015 to 417 in 2021, particularly among adults aged 50–79.

**Conclusions:** This study demonstrates that *H. pylori* eradication has already contributed to a 10.4% reduction in GC deaths in Japan by 2021, with annual expansion of primary prevention effects. This framework supports evidence-based evaluation of short-term reductions in GC deaths attributable to *H. pylori* eradication in high-prevalence settings.

## 1. Introduction

Gastric cancer (GC) remains a major public health concern in Japan, where it continues to be the third leading cause of cancer-related deaths [1].

*Helicobacter pylori* infection is the predominant etiologic factor [2], accounting for nearly all cases nationwide [3]. Although infection prevalence has declined across successive birth cohorts, Japan retains a unique epidemiologic structure in which older adults carry high lifetime exposure [4], shaping national GC trends for decades.

In 2013, Japan became the first country to introduce nationwide insurance coverage for *H. pylori* eradication therapy for chronic gastritis [5]. This policy led to a rapid expansion of eradication uptake and created a rare opportunity to evaluate the early, population-level impact of eradication in a real-world setting (Figure S1). Previous modeling work has demonstrated the cumulative lifetime impact of *H. pylori* eradication on GC deaths [6] and identified the optimal age for population-based implementation of eradication screening [7]. These findings highlight the importance of evaluating eradication strategies within a time-dependent, cohort-structured framework.

However, interpreting short-term GC deaths changes is inherently difficult because observed trends reflect multiple overlapping forces—population ageing, cumulative infection history [8], cohort replacement [9], and evolving clinical practice [10]— all of which vary by year. Conventional before-after comparisons or trend-based analyses cannot disentangle these structural determinants and therefore cannot isolate the causal contribution of eradication [11]. A counterfactual analysis is therefore needed to quantify the short-term divergence in GC deaths in the absence of eradication.

To address this complexity, the present study employs a two-layer analytic framework. First, a structured, time-dependent multilayer model reconstructs etiologic incidence by integrating age-specific biological hazard, duration-related exposure, and cohort-specific *H. pylori* prevalence. This modeling layer provides a biologically coherent representation of the causal pathway linking infection to GC. Second, a counterfactual analysis [12] compares observed GC deaths with an expected trajectory representing a scenario in which eradication uptake remained at pre-2013 levels. Together, these layers enables a population-level assessment of the early impact of eradication during its initial decade of nationwide implementation.

Using this combined modeling and counterfactual approach, the study aims to quantify the proportion of early reductions in GC deaths attributable to *H. pylori* eradication within the overall burden of GC deaths in Japan between 2013 and 2021. Clarifying these short-term population-level effects illustrates how primary prevention can begin to influence national cancer trends within a relatively brief period.

## 2. Methods

### 2.1 Study Design and Overview

We developed a structured, time-dependent multilayer model (Figure 1)[12, 13] to evaluate the population-level impact of Japan’s nationwide *H. pylori* eradication strategy implemented under the universal health insurance system.

**Figure 1.**
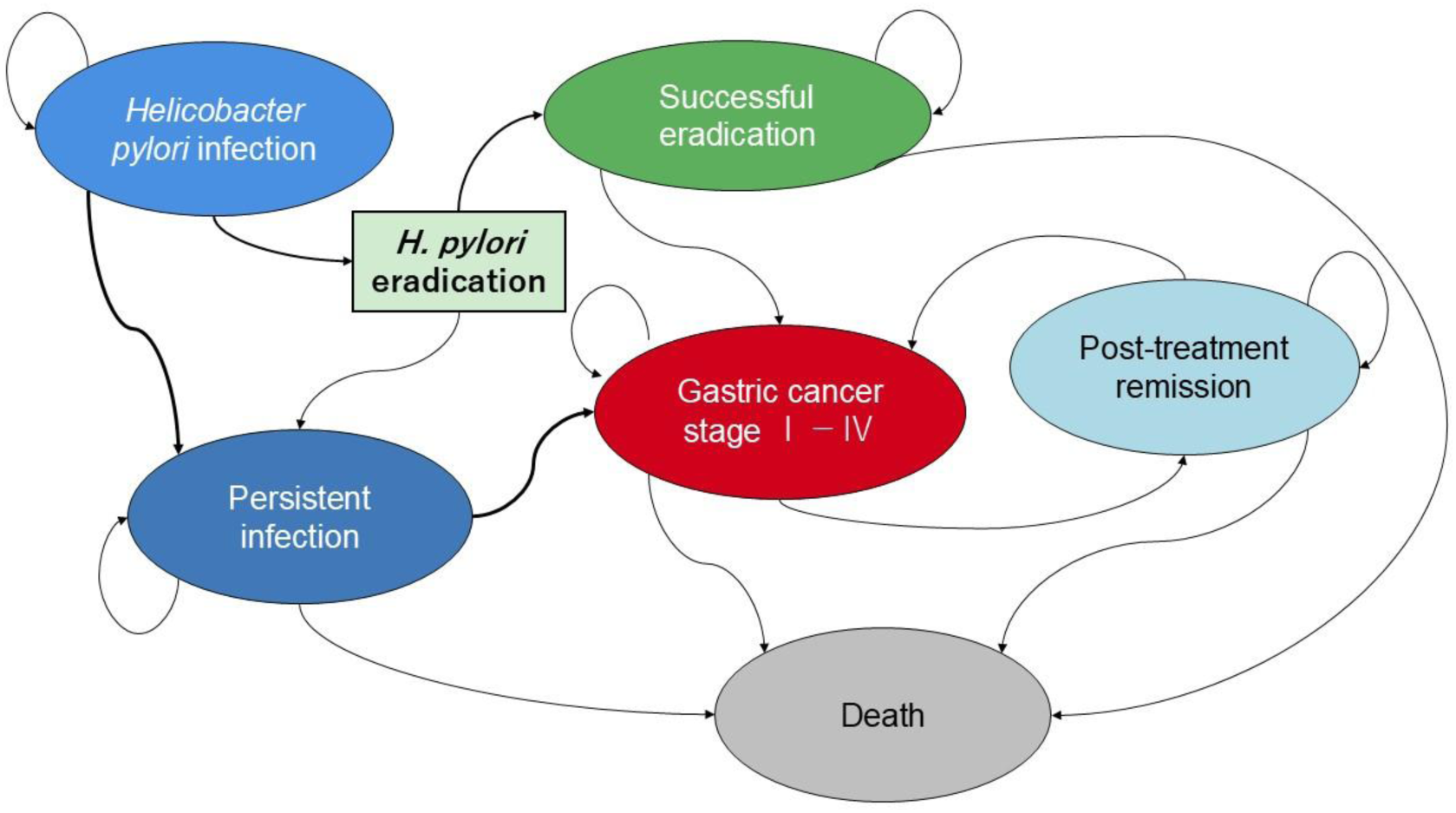
Clinical pathway and outcomes of *Helicobacter pylori* infection and eradication This diagram illustrates the progression and treatment outcomes associated with *H. pylori* infection and its potential link to gastric cancer. Infection may persist or be successfully eradicated through therapy. Persistent infection increases the risk of gastric cancer, which may lead to death or post-treatment remission. Successful eradication reduces gastric cancer risk. Arrows indicate possible transitions between health states, including recurrence after remission. Colors represent distinct clinical phases: blue for natural history, green for intervention, red for cancer progression, light blue for remission, and grey for death.

The analysis covered 2013–2021, corresponding to the period during which eradication uptake expanded rapidly following insurance coverage. Two scenarios were compared: the observed GC deaths reflecting real-world eradication uptake, and a counterfactual GC deaths trajectory representing expected GC deaths if eradication uptake had remained at pre-2013 levels. Comparing these scenarios isolates the contribution of eradication to short-term changes in GC deaths while preserving etiologic coherence with the established biology and epidemiology of *H. pylori*–related carcinogenesis.

### 2.2 Model Structure

#### 2.2.1 Multilayer GC incidence reconstruction

To capture the structural determinants of GC risk, we reconstructed age-specific GC incidence using a multiplicative formulation integrating four components [8,9]. First, baseline incidence was defined as the GC incidence at age 40 in 2021, obtained from the national cancer registry [1], and served as the reference scale. Second, an age-specific ageing factor was derived from 2021 incidence data to represent biological risk increases associated with ageing, independent of cohort exposure. Third, a duration-related historical exposure factor was estimated by comparing age-specific GC incidence in 2021 with incidence patterns from earlier high-burden periods; this factor captures cumulative historical exposure not explained by ageing alone. Fourth, age-and cohort-specific *H. pylori* prevalence was obtained from a systematic review and meta-regression of 170,752 individuals [4] and mapped to each age and calendar year.

The final incidence formulation was:

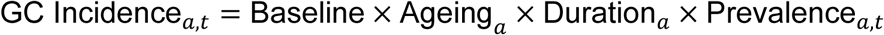

where *a* denotes age and *t* denotes calendar year.

This formulation incorporates cumulative infection history by combining age-related biological risk, duration of exposure, and cohort-specific *H. pylori* prevalence.

#### 2.2.2 GC deaths difference

GC deaths difference was defined as:

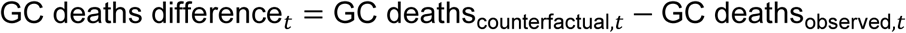

This difference reflects the divergence between expected and observed GC deaths in year *t*, capturing the overall reduction in GC deaths observed in real-world settings under existing screening practices.

#### 2.2.3 GC deaths prevented by eradication

GC deaths prevented were estimated using a structured state-transition (Markov) model comparing two strategies: continuation of standard GC screening and implementation of *H. pylori* eradication. This comparison yielded the number of GC deaths specifically attributable to eradication.

#### 2.2.4 Proportion of GC death reduction attributable to eradication

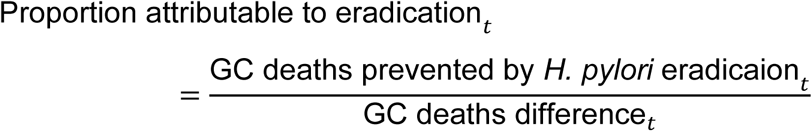

Here, GC deaths difference represents the observed–counterfactual divergence in year *t*, allowing estimation of the share of total GC death reduction explained by *H. pylori* eradication (Figure 2). This framework also enabled estimation of year-specific GC deaths prevented by eradication from 2013 to 2021.

**Figure 2.**
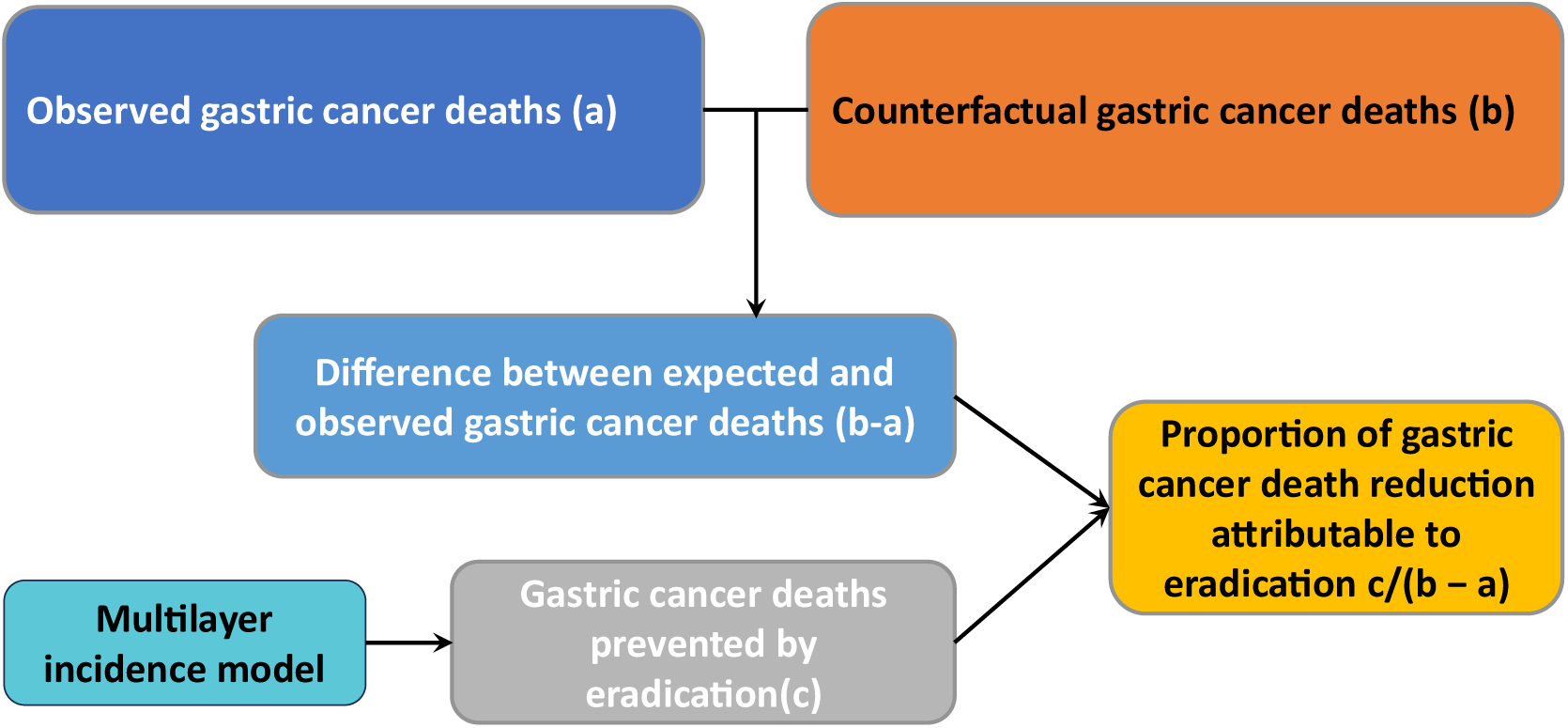
Conceptual framework for observed and counterfactual gastric cancer (GC) deaths and the estimated contribution of *H. pylori* eradication Observed GC deaths from 2013 to 2021 were compared with counterfactual GC deaths projected from mortality trends calibrated to national statistics from 1999 to 2012. The difference between expected and observed GC deaths (b − a) represents the real-world reduction in GC deaths under current screening practices. GC deaths prevented by eradication (c), estimated using a multilayer incidence model, together with the proportion of GC death reduction attributable to eradication (c / (b – a)), quantifies the contribution of *H. pylori* eradication to reductions in GC mortality.

### 2.3 Data inputs

Age-specific mortality, population structure, and GC incidence were obtained from national cancer statistics [1] and vital statistics [14] (Table 1). Annual eradication counts were derived from published literature [15] and non-public regional datasets (Table 2). Model parameters related to screening adherence [16], endoscopy accuracy [17], eradication compliance [18], eradication success [18], and GC risk reduction with eradication [19] were obtained from published studies (Table 1).

**Table 1.**
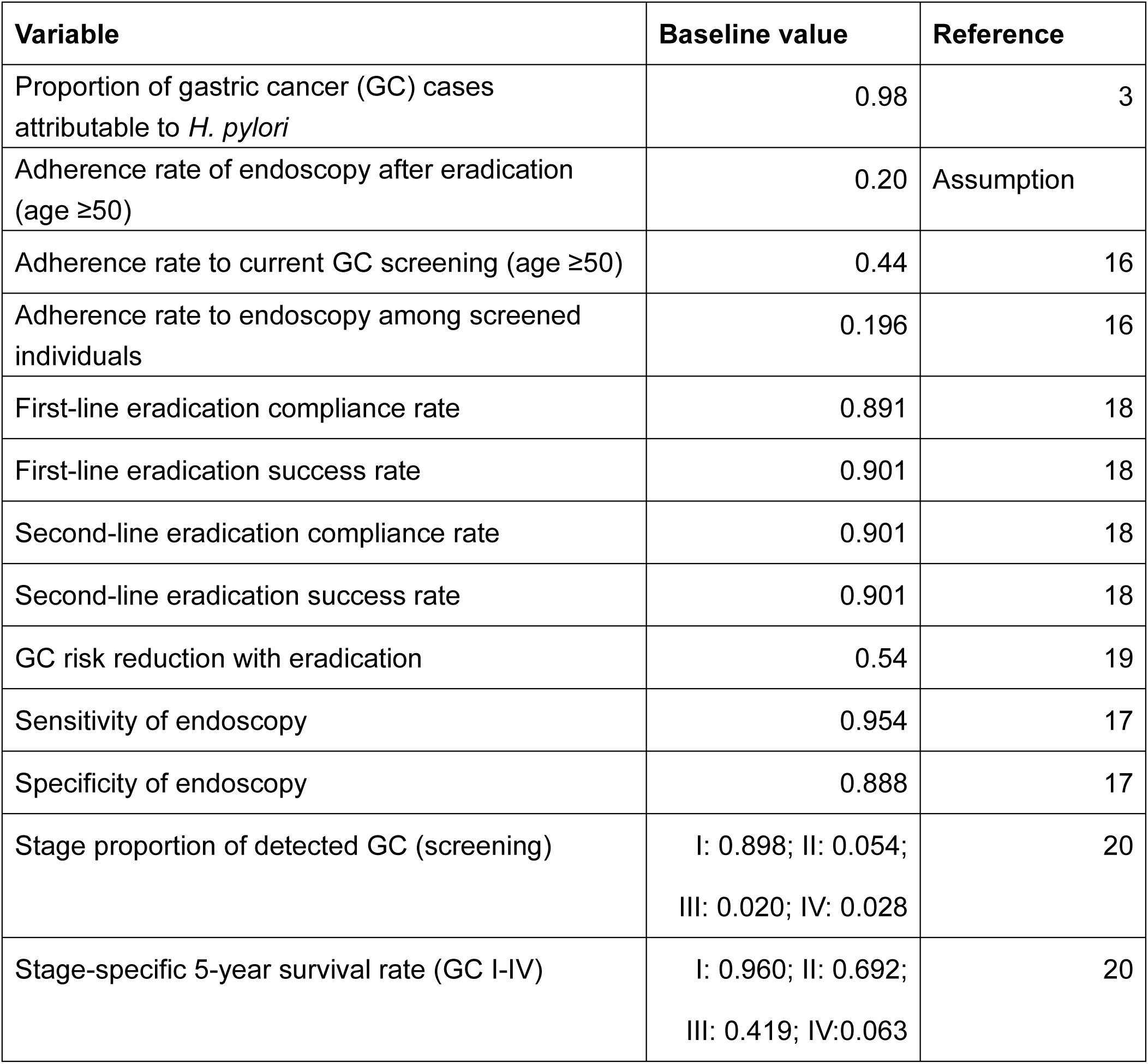
Key parameters used in the model.

**Table 2.** Annual differences between observed and counterfactual gastric cancer (GC) deaths and GC deaths prevented by *H. pylori* eradication, 2013–2021.

| **Year** | **Observed**<br>**GC deaths**<br>**(a)** | **Counterfactual**<br>**GC deaths (b)** | **Difference in**<br>**GC deaths**<br>**(b-a)** | **GC deaths**<br>**prevented by**<br>**eradication (c)** | **Proportion, %**<br>**(c/(b-a)) \*** |
| --- | --- | --- | --- | --- | --- |
| 2013 | 48,632 | 49,794 | 1,162 | 0 | 0 |
| 2014 | 47,904 | 49,717 | 1,813 | 0 | 0 |
| 2015 | 46,681 | 49,589 | 2,908 | 17 | 0.6 |
| 2016 | 45,546 | 49,310 | 3,764 | 47 | 1.3 |
| 2017 | 45,227 | 48,880 | 3,653 | 104 | 2.9 |
| 2018 | 44,192 | 48,300 | 4,108 | 194 | 4.7 |
| 2019 | 42,931 | 47,569 | 4,638 | 287 | 6.2 |
| 2020 | 42,319 | 46,687 | 4,368 | 361 | 8.3 |
| 2021 | 41,624 | 45,654 | 4,030 | 417 | 10.4 |
| Total | 405,056 | 435,500 | 30,444 | 1,427 | 4.7 |

\*Proportion of the observed–counterfactual GC death difference attributable to eradication.

Epidemiologic parameters related to GC progression were derived from national statistics [20] and from prior long-term modeling work evaluating the cumulative lifetime impact of eradication on GC deaths [6]. Age-specific stage distributions for screened GC were incorporated using national cancer statistics [20], and are consistent with evidence showing that younger patients typically present with more advanced disease and aggressive tumor biology [21,22] (Table S1).

All data sources were harmonized to produce internally consistent age-and year-specific inputs. All modeling analyses were conducted using TreeAge Pro 2025 (TreeAge Software, Williamstown, MA).

## 3. Results

### 3.1 Trends and divergence between observed and counterfactual GC deaths

Annual trends in the number of individuals undergoing *H. pylori* eradication are shown in Figure 3A, demonstrating a rapid increase following the introduction of insurance coverage in 2013, followed by a gradual decline and recent plateau. Observed GC deaths decreased steadily from 48,632 in 2013 to 41,624 in 2021 (Table 2, Figure 3B).

**Figure 3.**
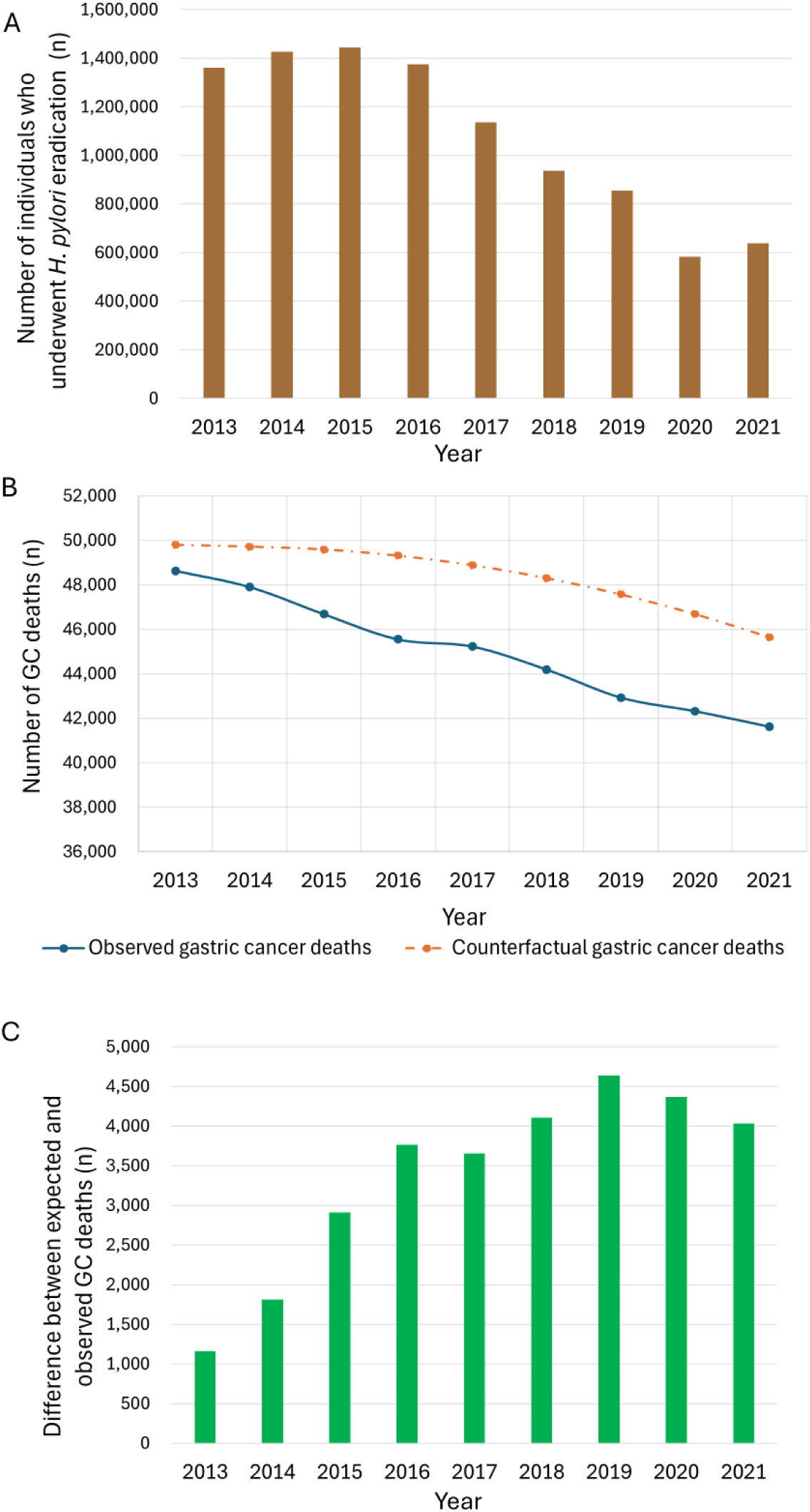
Temporal trends in the number of individuals who underwent *H. pylori* eradication and gastric cancer (GC) deaths in Japan, 2013–2021 A. Number of individuals who underwent *H. pylori* eradication B. Number of observed and counterfactual GC deaths C. Difference between expected and observed GC deaths

In contrast, the counterfactual GC deaths trajectory—representing expected GC deaths if eradication uptake had remained at pre-2013 levels—declined more slowly, from 49,794 in 2013 to 45,654 in 2021 (Table 2, Figure 3B).

The divergence between observed and counterfactual GC deaths widened consistently over time, increasing from 1,162 deaths in 2013 to 4,030 in 2021 (Table 2, Figure 3C). This widening gap reflects the cumulative impact of expanded eradication uptake on national mortality trends.

### 3.2 GC deaths prevented by eradication

Model-based estimates indicated that the number of GC deaths prevented by eradication increased gradually throughout the study period. GC deaths prevented rose from 17 in 2015—the first year in which measurable effects of eradication appeared—to 417 in 2021, with a cumulative total of 1,427 prevented during 2013–2021 (Table 2, Figure 4A).

**Figure 4.**
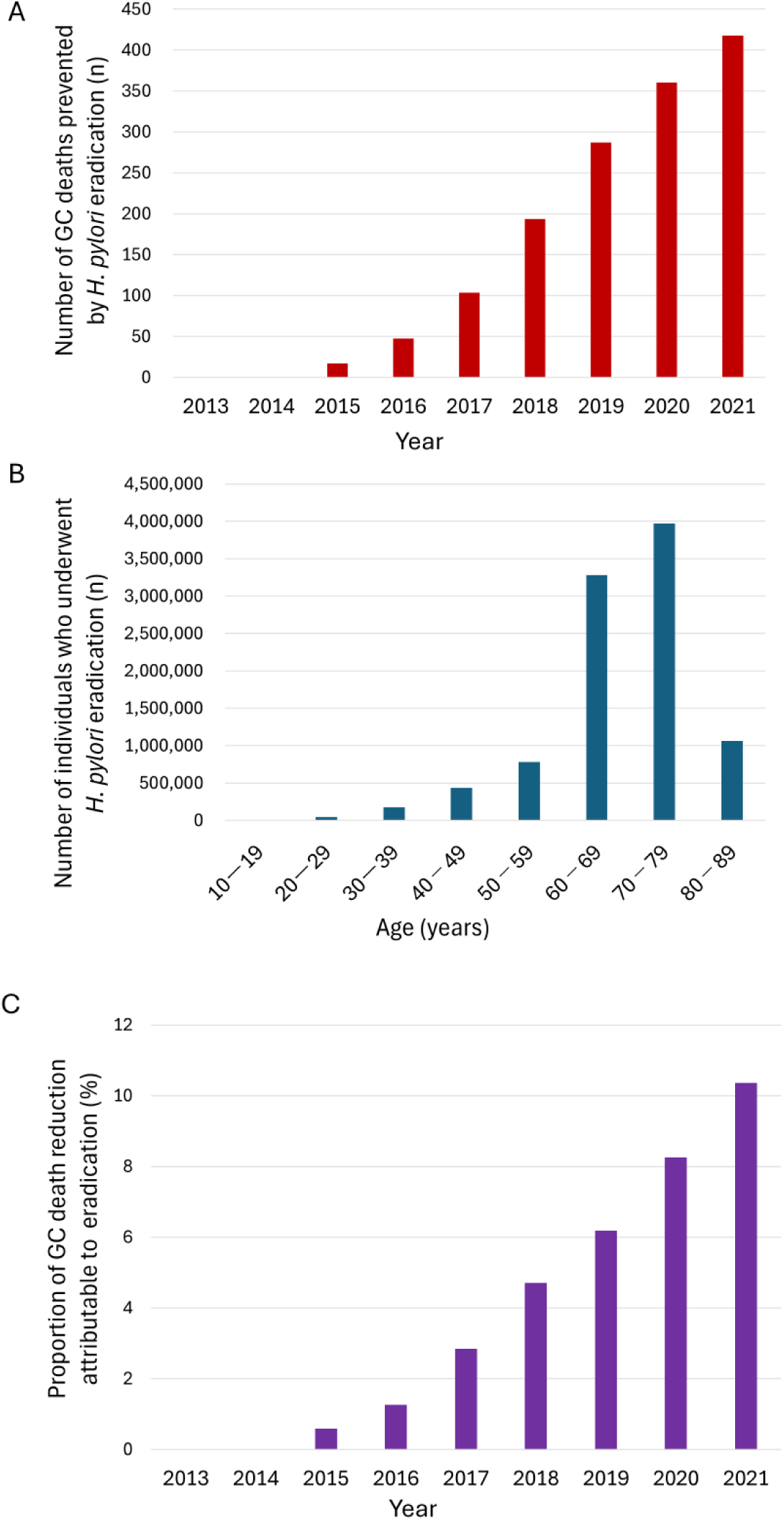
Gastric cancer (GC) deaths prevented by *H. pylori* eradication, 2013–2021 A. Number of GC deaths prevented by *H. pylori* eradication B. Number of individuals who underwent *H. pylori* eradication by age group C. Proportion of GC death reduction attributable to eradication

The largest annual reductions in GC deaths were observed among adults aged 50–79, consistent with both the age distribution of eradication uptake and the cohort-specific infection histories that shape GC risk (Figure 4B).

### 3.3 Proportion of GC death reduction attributable to eradication

The proportion of the observed–counterfactual GC death difference attributable to eradication increased steadily over time. This proportion rose from 0.6% in 2015 to 10.4% in 2021 (Table 2, Figure 4C).

This upward trend indicates that eradication has become an increasingly important contributor to Japan’s declining GC deaths, even within the relatively short nine-year period following nationwide insurance coverage.

### 3.4 Summary of early population-level impact

Taken together, these findings demonstrate that nationwide expansion of *H. pylori* eradication has already produced measurable reductions in gastric cancer mortality at the population level. The observed decline in deaths has consistently outpaced the counterfactual trajectory throughout the post-2013 period, and the contribution of eradication to this divergence has increased steadily over time. Even within the first nine years of insurance coverage, eradication has emerged as a meaningful driver of Japan’s declining GC mortality, indicating that its population-level benefits are both early and progressively increasing.

## 4. Discussion

### 4.1 Interpretation of findings

By integrating a structured incidence reconstruction with a counterfactual GC deaths trajectory, this study clarifies the early population-level impact of Japan’s nationwide *H. pylori* eradication strategy. Mortality declined more rapidly than expected in 2015, and the divergence between observed and counterfactual GC deaths widened steadily through 2021. These findings indicate that eradication has already produced measurable reductions in GC deaths within the first decade of implementation, despite the long latency of gastric carcinogenesis.

The strongest effects were observed among adults aged 50–79, reflecting both the age distribution of eradication uptake and the cumulative infection histories that shape GC risk.

### 4.2 Structural epidemiologic implications

This study demonstrates the value of a two-layer analytic framework that combines etiologic modeling with counterfactual evaluation. The multilayer incidence reconstruction captures the biological and historical determinants of GC risk—age-specific hazard, duration-related exposure, and cohort-specific infection prevalence—providing a coherent representation of the causal pathway linking *H. pylori* infection to cancer. The counterfactual layer isolates the contribution of eradication from background trends driven by ageing, cohort replacement, and long-term declines in infection prevalence. Together, these components enable a causal interpretation of short-term mortality changes that conventional trend-based or before–after analyses cannot achieve.

### 4.3 Policy relevance

Japan’s experience provides real-world evidence that population-based *H. pylori* eradication can produce detectable reductions in GC deaths within a relatively short timeframe. The alignment between eradication volume, early mortality reductions, and modeled GC deaths prevented supports the continued expansion of eradication as a primary prevention strategy. These findings are particularly relevant for high-prevalence countries where GC remains a major cause of cancer mortality. As younger cohorts with lower infection prevalence gradually replace older high-risk cohorts, the relative contribution of eradication to national mortality trends may increase further. Integrating eradication with risk-based screening strategies may accelerate progress toward reducing GC burden.

### 4.4 Comparison with conventional modeling and forecasting approaches

Unlike conventional natural history models that rely on unobservable transition parameters, the present framework reconstructs incidence directly from etiologic determinants. This etiology-based approach avoids assumptions about latent disease states and provides a transparent representation of the causal structure underlying GC risk. Forecasting-based approaches, including ARIMA and interrupted time-series regression [23], are valuable for projecting future trends but cannot isolate the causal impact of specific policy actions because they extrapolate past patterns without reconstructing counterfactual disease dynamics. In contrast, the combined modeling and counterfactual design used here quantifies the short-term mortality reduction attributable specifically to real-world eradication efforts.

### 4.5 Strengths and limitations

Strengths of this study include the use of a counterfactual framework explicitly aligned with real-world policy implementation, which enabled estimation of mortality trends under a plausible no-eradication scenario. Additional strengths include reconstruction of GC incidence based on etiologic determinants and the integration of age-specific *H. pylori* prevalence, case fatality, and eradication uptake. This approach allowed quantification of both the absolute number of GC deaths prevented and the proportion of national mortality decline attributable to eradication.

Limitations include reliance on published case-fatality estimates and assumptions regarding the stability of background risk factors in the counterfactual scenario. The analysis focuses on short-term effects and does not project long-term outcomes, which may differ as younger cohorts with lower *H. pylori* prevalence age into higher-risk periods. However, our previous work demonstrated the greater long-term effects of eradication [6]. Temporal changes in non–*H. pylori* risk factors, such as high salt intake, smoking, obesity, and excess alcohol consumption, were not explicitly modeled, although their short-term influence is likely modest compared with the impact of eradication.

### 4.6 Implications

This structured counterfactual analysis demonstrates that population-based *H. pylori* eradication can produce measurable reductions in GC deaths within a decade. The findings highlight the importance of integrating real-world implementation patterns, etiologic determinants, and causal evaluation frameworks when assessing primary prevention strategies. More broadly, this study provides a complementary perspective to global analyses by emphasizing the role of historical implementation context and short-term dynamics in shaping national cancer trends.

## 5. Conclusion

This study provides clear evidence that population-based *H. pylori* eradication has already contributed to measurable reductions in GC deaths in Japan. By combining a structured etiologic modeling framework with a counterfactual evaluation of expected mortality in the absence of expanded eradication, the analysis clarifies how eradication is shaping national GC deaths trends during the early phase of its implementation. These findings demonstrate that primary prevention can yield detectable population-level benefits within a decade, even for a cancer with a long natural history. Japan’s experience offers real-world evidence of how systematic eradication can alter short-term national cancer trajectories and provides a framework for evaluating early implementation effects in high-prevalence settings.

## Data Availability

All aggregated national statistics analyzed in this study are publicly available from official sources. Some regional datasets were provided under restricted data use agreements and are not available for public release.

https://ganjoho.jp/reg_stat/statistics/stat/cancer/5_stomach.html

https://www.mhlw.go.jp/toukei/saikin/hw/k-tyosa/k-tyosa22/index.html.

https://ganjoho.jp/public/qa_links/report/statistics/2025_en.html

https://www.mhlw.go.jp/english/database/db-hw/vs01.html

## Acknowledgments

None.

## Ethics Statement

This study used publicly available, aggregated national statistics and did not involve human subjects; therefore, ethical approval was not required.

## Author Contributions

The author conceptualized the study, developed the model, conducted the analysis, validated the model, interpreted the results, drafted the manuscript, and revised the final version.

