## Supplementary Material for "Early Population-Level Impact of *Helicobacter pylori* Eradication on Gastric Cancer Deaths in Japan: A Counterfactual Analysis of Short-Term Divergence"

**Table S1.** Age-specific stage distribution of gastric cancer detected without screening

| Age group | Stage I | Stage II | Stage III | Stage IV |
|---|---|---|---|---|
| 10–19 | 0.224 | 0.074 | 0.137 | 0.634 |
| 20–29 | 0.342 | 0.068 | 0.124 | 0.466 |
| 30–39 | 0.500 | 0.094 | 0.104 | 0.302 |
| 40–49 | 0.577 | 0.098 | 0.092 | 0.233 |
| 50–59 | 0.631 | 0.084 | 0.094 | 0.191 |
| 60–69 | 0.637 | 0.077 | 0.098 | 0.188 |
| 70–79 | 0.659 | 0.077 | 0.090 | 0.174 |
| 80–89 | 0.613 | 0.093 | 0.096 | 0.198 |

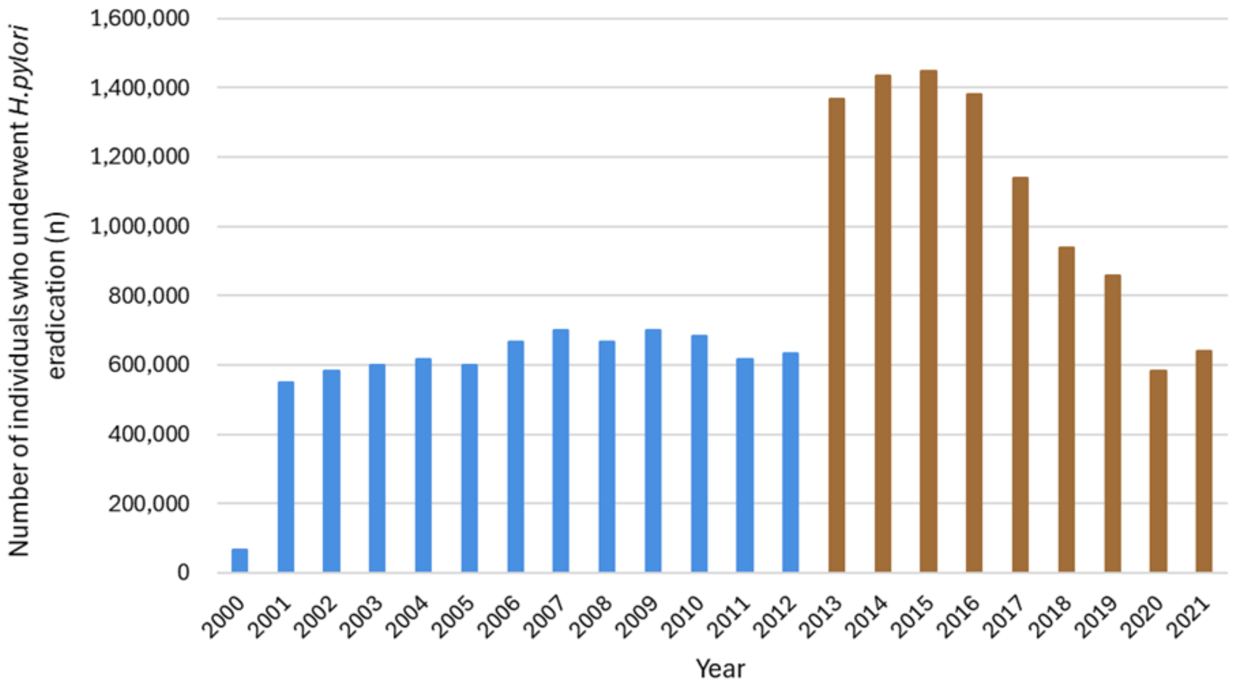


**Figure S1.** Temporal trend in the number of individuals who underwent *Helicobacter pylori* eradication in Japan, 2000–2021

Eradication therapy for peptic ulcer disease has been covered by national insurance since 2000, but the number of individuals receiving eradication remained modest until 2012. The number increased sharply after insurance coverage was expanded to include chronic gastritis in 2013.
